# Post-Acute Rehabilitation Placement After Acute Ischemic Stroke Is Associated With Non-Clinical Factors Despite Similar Clinical Profiles

**DOI:** 10.64898/2026.05.08.26352775

**Authors:** Heather Hayes, Chong Zhang, Siqi Xiang, Bradley Smith, Perry Williams, Angela P Presson, Maggie French

## Abstract

**Background:** Discharge destination after acute ischemic stroke has implications for functional recovery and healthcare costs. Individuals discharged to inpatient rehabilitation facilities (IRFs) achieve better outcomes than those discharged to skilled nursing facilities (SNFs); however, many patients discharged to IRFs and SNFs have similar clinical profiles. We examined non-clinical factors associated with discharge location after acute ischemic stroke.

**Methods:** Population: 236 adults hospitalized with acute ischemic stroke, living independently in the community prior to admission, and discharged to either an IRF (n=171) or SNF (n=65). Clinical variables: NIHSS, Charlson Comorbidity Index (CCI), acute care length of stay (LOS), functional status (AM-PAC “6-Clicks”), and neglect. Non-clinical variables: age, sex, race, marital status, insurance, home layout, living status, and available assistance. Associations with discharge location were evaluated using univariable and multivariable logistic regression and reported as odds ratios (OR) with 95% confidence intervals (CI).

**Results:** Individuals discharged to IRFs were younger, more likely to cohabitate, and had shorter LOS than those discharged to SNFs. Functional status (AM-PAC) and comorbidity burden (CCI) did not differ significantly between groups despite differences in discharge destination. In univariable models, younger age, cohabitating marital status, living with family, available assistance, shorter LOS, private insurance, and higher NIHSS were associated with greater odds of IRF discharge. In multivariable analysis, younger age (OR 0.94, 95% CI 0.91–0.98), cohabitating marital status (OR 2.46, 95% CI 1.13–5.48), and shorter LOS (OR 0.88, 95% CI 0.82–0.93) remained independently associated with IRF discharge.

**Conclusions:** Individuals with comparable pre-stroke independence and similar clinical severity, discharge to IRF versus SNF was independently associated with non-clinical factors; age, marital status, and LOS, whereas stroke severity and functional status were not significant predictors. These findings underscore the importance of evidence-informed discharge criteria integrating clinical indicators and social context to support equitable access to intensive rehabilitation after stroke.

## Introduction

Stroke is a leading cause of long-term disability in the United States, and the number of survivors requiring post-acute rehabilitation care (PARC) is expected to increase in the coming decades.^1^ The early recovery period, particularly the first 90 days post-stroke, is critical, as access to intensive rehabilitation during this window can optimize functional recovery and reduce long-term disability.^2^ Discharge planning from acute hospitalization, specifically whether individuals post-stroke transition to an inpatient rehabilitation facility (IRF) or skilled nursing facility (SNF), is a pivotal decision with profound functional and economic consequences, as these settings differ substantially in rehabilitation intensity during the critical early post-stroke recovery period.^3^

Clinical guidelines recommend IRF care over SNF care for individuals post-stroke, as IRF treatment is consistently associated with greater functional improvement, higher rates of community discharge, lower readmission, and lower mortality.^4–6^ Despite these recommendations, a larger proportion of stroke survivors in the United States are discharged to SNFs rather than IRFs.^4^ This is at least in part due to policy guidelines for determining discharge to an IRF versus SNF. Specifically, IRFs require that individuals are able to tolerate intensive multidisciplinary therapy (≥3 hours/day, 5–6 days/week) and have the capacity for measurable functional improvement and community reintegration, whereas SNFs provide less intensive therapy coupled with nursing care.^4^ However, implementation of these recommendations varies considerably across institutions and regions, resulting in discharge practices that are influenced by non-clinical, hospital-level, and sociodemographic factors in addition to patient impairment severity.^3,6–8^

Although policy requirements partially shape discharge placement, substantial variability in referral patterns persists even among individuals with similar stroke severity and functional impairment. The factors driving these differences remain incompletely understood. Prior research has identified a range of clinical factors^9^ (e.g. stroke severity, comorbidities, and functional status) and non-clinical factors; 1) demographics^10–13^ (age, sex, race) and 2) contextual factors^6,14^ (e.g. marital status, insurance, living arrangement, home layout, and available assistance). Findings, however, have been inconsistent.

Importantly, prior studies suggest that individuals with comparable stroke severity and functional impairment are frequently discharged to different post-acute settings (IRF vs SNF), despite these characteristics being central determinants of rehabilitation intensity and recovery potential.^11^ Such variation raises concern that discharge placement may be influenced not only by measurable clinical indicators, but also by demographic and contextual characteristics unrelated to neurologic impairment alone.

Clarifying the relative contributions of clinical and non-clinical factors to discharge location is essential for ensuring equitable and evidence-based access to post-acute rehabilitation. Therefore, the aim of this study was to examine clinical and non-clinical (demographics and contextual) factors associated with discharge location to either an IRF or SNF among individuals hospitalized with acute ischemic stroke who were independent and community-dwelling prior to stroke onset. We hypothesized that non-clinical factors would demonstrate independent associations with discharge destination, even after accounting for clinical indicators of stroke severity.

## Methods

### Study design and cohort selection

This study was performed at the University of Utah hospital, Salt Lake City, UT, USA. The IRF cohort was collected prospectively and included individuals admitted to the acute care hospital with a confirmed diagnosis of ischemic stroke verified by imaging (CT or MRI) and provided consent (IRB_00114762). Data for the SNF cohort were collected retrospectively from the Enterprise Data Warehouse at the University of Utah (IRB_001171867) spanning January 1, 2020 to July 31, 2024. The SNF cohort was only retrospective due to limited prospective data collection during 2020 – 2021 with the COVID-19 pandemic. The SNF sample included individuals admitted into the acute care hospital under Neurology or Internal Medicine with a diagnosis of ischemic stroke (ICD-9-CM codes 433-434, 436; ICD-10 code I63.9) listed in the top three provider billing diagnoses.

For both cohorts, we included adult individuals (≥18 years) hospitalized with an acute ischemic stroke who were discharged from acute care to either an IRF or SNF. We included only individuals with a pre-stroke level of function (PLOF) characterized as independent and who were living in the community prior to their stroke (i.e., not residing in long-term care or other institutional settings), which was taken from chart review as part of routine questioning by the physical and/or occupational therapist and recorded in a discrete field.

For both cohorts, we excluded individuals that were incarcerated, and those missing an NIHSS assessment. The final analytic sample included 236 individuals, 171 discharged to an IRF and 65 to a SNF (see enrollment diagram, Supplement 1). Data collection spanned from January 1, 2020 to July 31, 2024 for both the IRF (prospective data) and SNF (retrospective data) cohorts.

### Variables and measures

The primary outcome was discharge location from the acute hospital (IRF vs. SNF). Clinical factors were defined as variables reflecting stroke severity, comorbidity burden, and functional status during acute hospitalization. The clinical factors represent the patient’s clinical presentation at the time of admission and includes 7 variables; 1) National Institutes of Health Stroke Scale (NIHSS); 2) Charlson Comorbidity Index (CCI); 3) acute care clinical length of stay (LOS); (4-6) functional presentation based on the three domains of the Activity Measure for Post-Acute Care (AM-PAC) scores: Basic Mobility (BM), Daily Activities (DA), and Applied Cognition (AC); and (7) presence of neglect or not [present, absent or missing]. The NIHSS is a rating of stroke severity, evaluating 11 areas and scores can range from 0 (no stroke) to 42 (severe stroke symptoms). Neglect was identified from the NIHSS (item 10) and clinical chart reviews. The AM-PAC “6-Clicks” was taken from chart review and is required to be performed by Physical and Occupational Therapy services. The raw score ranges from 6 to 24, with higher scores indicating greater independence.

Non-clinical factors were defined as demographic and contextual characteristics not directly reflective of neurologic impairment or functional status. The non-clinical factors that we examined included 3 demographic variables and 5 contextual variables.

Demographic variables included: 1) age [years]; 2) sex [female or male]; 3) race [white or other/unknown]. Demographic variables were obtained from discrete fields in the chart for both cohorts. The contextual variables included: 1) marital status; 2) primary insurance; 3) home layout; 4) available assistance; 5) who patient lives with. Marital status was coded as alone [if divorced, separated, single, or widowed] or cohabitates [life partner or married] or missing. Primary insurance type was coded as Private, Medicare, Medicare Advantage (MA), Medicaid, or none. Home layout data was defined as structural barriers present [stairs in the living setting, stairs to get into home, narrow doors, and/or limited space for equipment] or not present or missing. Available assistance was coded as yes, if they required full-time / part-time physical assistance or full-time / part-time supervision. Lives with status was coded as individuals living in the community alone [individuals lived alone] or with family/other [paid helper, living with family, or unknown] or missing. Contextual variables were obtained from discrete fields in the chart for both cohorts.

## Statistical Analysis

Descriptive statistics were calculated for all variables stratified by discharge location. Continuous variables were compared using the Wilcoxon rank-sum test, and categorical variables using Chi-squared or Fisher’s exact tests, as appropriate.

Univariable logistic regression was used to assess the unadjusted association between each factor and discharge location. Variables with *p* < 0.10 were considered for inclusion in a multivariable logistic regression model. Collinearity among variables was evaluated using variance inflation factors (VIFs) with GVIF^1/2df^>2 considered suggestive of multicollinearity. Because the number of variables was limited by the number of individuals, only one variable was selected from related and correlated variables, prioritizing conceptual relevance and completeness of data. Missing data were handled by listwise deletion. Model results were reported as odds ratios (OR) with 95% confidence intervals (CI) and p-values. All analyses were conducted in R v4.3.127 (R Core Team, 2023) using two-tailed tests. Statistical significance was evaluated at the 0.05 level.

## Results

Of the 236 individuals included in the analysis, 171 (72%) were discharged to an IRF and 65 (28%) to a SNF. Compared with individuals discharged to a SNF, those discharged to an IRF were younger (mean age 64 vs 74 years, *p* < 0.001), more likely to cohabitate (*p* = 0.006), had shorter LOS (mean 7.8 vs 13 days, *p* < 0.001), and has more severe strokes per the NIHSS (mean 10.0 vs 7.7 points, p = 0.006). Additional group differences were observed in insurance type, barriers in home layout, living status, AM-PAC Basic Mobility and Applied Cognition scores (**Table 1**).

**Table 1.**
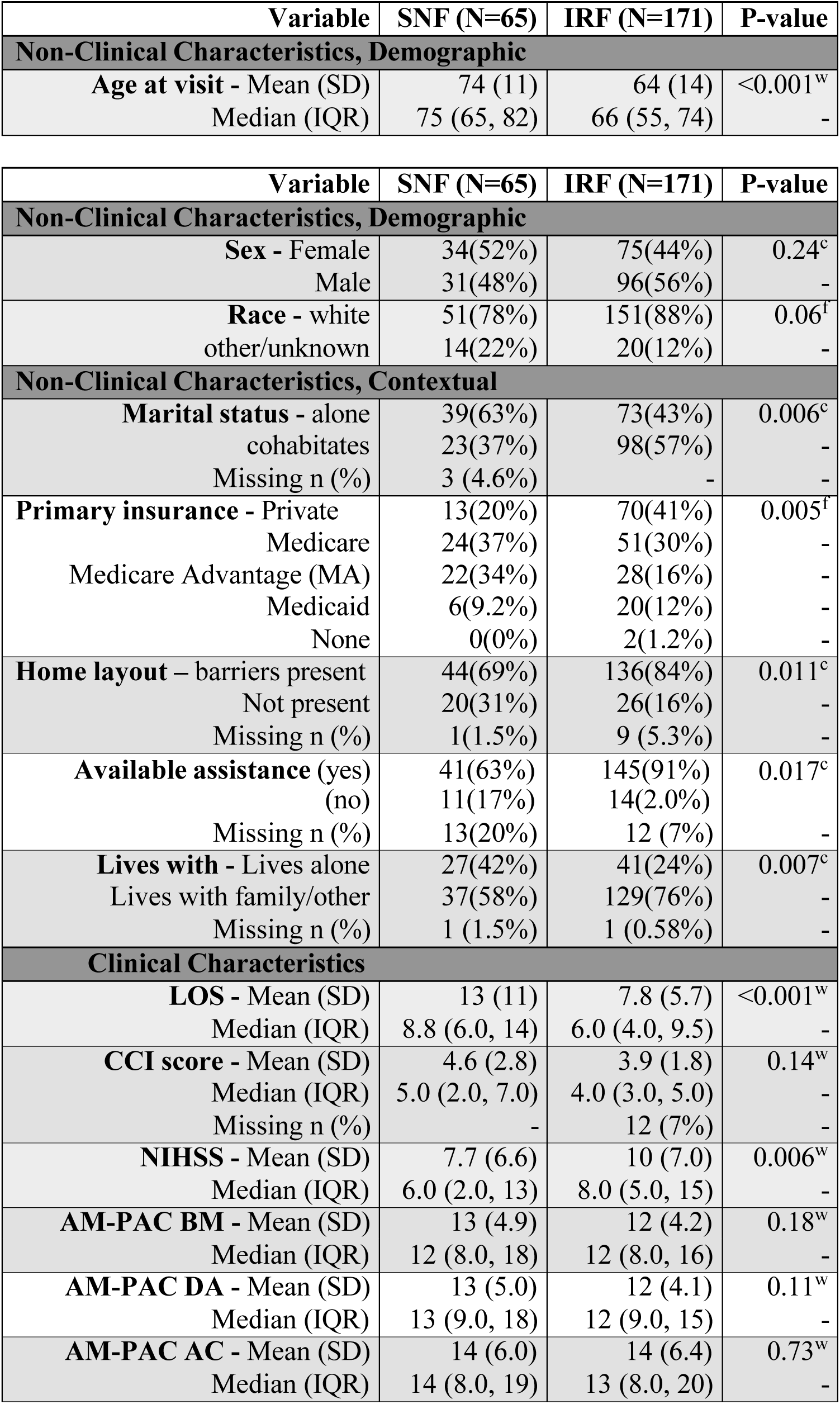
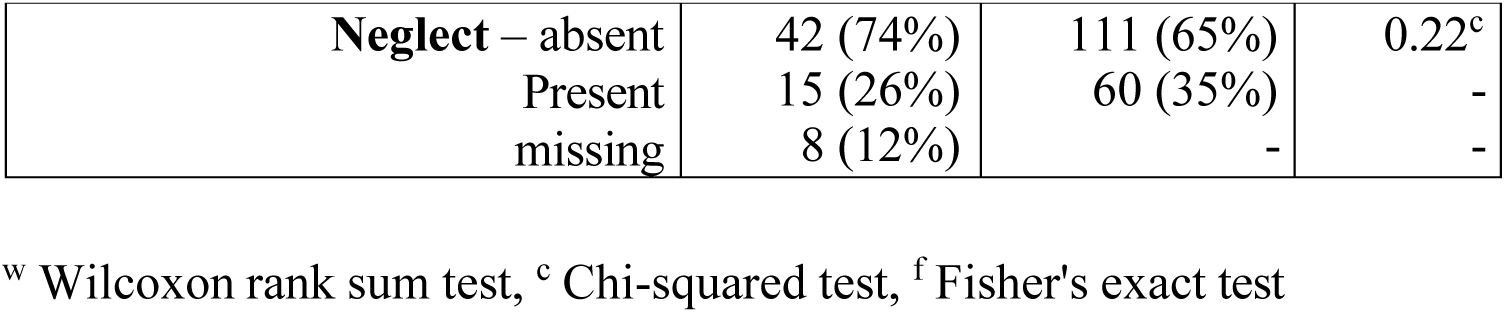
Descriptive summary of individuals post-stroke.

In univariable logistic regression (Table 2), several non-clinical (demographic and contextual) factors were associated with increased odds of discharge to an IRF, including cohabitating marital status (OR 2.28, 95% CI 1.26–4.19), living with family (OR 2.30, 95% CI 1.25–4.22), available assistance (OR 2.78, 95% CI 1.15–6.58), and private insurance (OR 2.53 vs. Medicare, 95% CI 1.20–5.58). Higher NIHSS was also associated with increased odds of IRF discharge (OR 1.06, 95% CI 1.01–1.11). In contrast, increasing age (OR 0.94 per year, 95% CI 0.92–0.97) and longer clinical LOS (OR 0.92 per day, 95% CI 0.88–0.96) were associated with reduced odds of discharge to an IRF. Since three contextual variables; 1) marital status, 2) available assistance, and 3) who individuals lived with were related, we chose marital status as the representative of these three variables for our multivariable model. In the final multivariable logistic regression model (N = 211 [SNF=61, IRF=150]; **Table 2**), three variables remained independently associated with IRF discharge: 1) increasing age, OR 0.94 (95% CI 0.91–0.98), *p* = 0.002; 2) cohabitating marital status, OR 2.46 (95% CI 1.13–5.48), *p* = 0.025; and 3) LOS: OR 0.88 (95% CI 0.82–0.93), *p* < 0.001.

**Table 2.** Univariable and Logistic Regression models predicting IRF discharge.

| Variable | OR (univariable) | P-value | OR (multivariable)** | P-value |
| --- | --- | --- | --- | --- |
| **Non-Clinical Characteristics, Demographic** |  |  |  |  |
| **Age** | 0.94 (0.92, 0.97) | <0.001 | 0.94 (0.91, 0.98) | 0.002 |
| **Sex (Male)** | 1.40 (0.79, 2.50) | 0.25 | - | - |
| **Race (Other/unknown)** | 0.48 (0.23, 1.04) | 0.06 | - | - |
| **Non-Clinical Characteristics, Contextual** |  |  |  |  |
| **Marital Status** (cohabitates vs. alone) | 2.28 (1.26, 4.19) | 0.007 | 2.46 (1.13, 5.48) | 0.025 |
| **Primary Insurance\*** |  |  |  |  |
| (Private vs Medicare) | 2.53 (1.20, 5.58) | 0.017 | 1.26 (0.45, 4.23) | 0.59 |
| (MA vs Medicare) | 0.60 (0.28, 1.26) | 0.17 | 0.61 (0.24, 1.51) | 0.29 |
| (Medicaid vs Medicare) | 1.57 (0.58, 4.74) | 0.39 | 0.81 (0.17, 4.27) | 0.80 |
| **Home Layout** (not present) | 0.42 (0.21, 0.83) | 0.012 | 0.66 (0.28, 1.59) | 0.35 |
| **Available Assistance** (Yes) | 2.78 (1.15, 6.58) | 0.020 | - | - |
| **Lives with** (family/other) | 2.30 (1.25, 4.22) | 0.007 | - | - |
| **Clinical Characteristics** |  |  |  |  |
| **LOS** | 0.92 (0.88, 0.96) | <0.001 | 0.88 (0.82, 0.93) | <0.001 |
| **CCI Score** | 0.87 (0.76, 0.99) | 0.043 | 0.87 (0.74, 1.01) | 0.08 |
| **NIHSS** | 1.06 (1.01, 1.11) | 0.014 | 1.03 (0.97, 1.10) | 0.27 |
| **AM-PAC BM** | 0.95 (0.89, 1.02) | 0.14 | - | - |
| **AM-PAC DA** | 0.94 (0.88, 1.01) | 0.09 | 0.93 (0.84, 1.03) | 0.18 |
| **AM-PAC AC** | 1.01 (0.96, 1.06) | 0.74 | - | - |
| **Neglect** (present) | 1.51 (0.79, 3.02) | 0.22 | - | - |

\*For two individuals with no insurance, we assigned NA to their insurance values so that they were excluded from the evaluation of insurance types.
\*\*Sample size for the multivariable regression model: N=211 (SNF=61, IRF=150).

Insurance type, comorbidity burden, NIHSS, and AM-PAC DA scores did not remain significant after adjustment.

## Discussion

In this study of individuals hospitalized with acute ischemic stroke who were living independently in the community prior to hospitalization, we examined clinical and non-clinical (demographic and contextual) factors associated with discharge to either an IRF or a SNF. In this cohort of previously independent, community-dwelling individuals hospitalized with acute ischemic stroke, discharge to IRF versus SNF was independently associated with non-clinical factors, particularly age and marital status, despite comparable measured clinical severity and functional status between groups. These findings support our hypothesis that non-clinical factors demonstrate independent associations with discharge placement decisions.

First, younger individuals were significantly more likely to be discharged to IRFs even after adjustment for stroke severity. Although age is consistently associated with poorer post-stroke outcomes was,^6,7,12,15^ these difference may partly reflect unequal access to high-intensity rehabilitation, given the documented benefits of IRF care.^3,4,16,17^ Older adults are more frequently discharged to SNFs, where recovery trajectories are typically less favorable.³ Thus, age may reflect not only biological variation in recovery potential, but also structural differences in post-acute care allocation. In practice, older age may be implicitly interpreted as reduced ability to tolerate intensive rehabilitation despite limited evidence that chronological age alone accurately reflects rehabilitation capacity or potential benefit from high-intensity therapy.

Second, marital status was independently associated with IRF discharge, suggesting that perceived availability of home support influences post-acute placement decisions. Although caregiver support may facilitate successful community reintegration, reliance on marital or living status as a proxy for rehabilitation suitability may inadvertently influence access to intensive rehabilitation services.

Third, shorter acute care LOS was independently associated with IRF discharge. Although prolonged hospitalization may reflect greater medical complexity, measured comorbidity burden and functional status were similar between groups. LOS may therefore capture broader clinical and logistical processes, including delays in post-acute placement, institutional workflows, or clinician perceptions of rehabilitation readiness. This finding suggests that hospital-course factors may indirectly shape post-acute rehabilitation allocation independent of measurable stroke severity.

Notably, functional status as measured by the AM-PAC did not differ between groups and was not independently associated with discharge location. These findings indicate that measurable functional impairment did not account for discharge placement in this cohort. In contrast, non-clinical contextual factors (age and marital status) were independently associated with referral to IRF versus SNF. Although insurance demonstrated associations in unadjusted models, consistent with prior research,^6^ it did not remain significant after adjustment, possibly reflecting limited statistical power.

Collectively, these findings suggest that post-acute rehabilitation placement after stroke may not consistently align with measurable indicators of rehabilitation need alone. Despite similar functional status and comorbidity burden, demographic and contextual characteristics remained independently associated with discharge destination. Although IRF admission emphasize medical stability, tolerance for intensive interdisciplinary rehabilitation, and anticipated functional improvement,^18^ how these criteria are operationalized in clinical practice may vary across institutions and clinicians. Greater transparency and evidence-informed frameworks for discharge planning may help ensure equitable access to intensive rehabilitation services.

## Limitations

This study has several limitations. First, all data was captured from within a single health system, which may limit generalizability to other regions or practice environments. Second, racial differences in discharge destination could not be robustly evaluated because our sample was predominantly White, reflecting regional demographics. Prior research has identified racial disparities in post-acute placement, warranting further investigation in more diverse cohorts. Third, the SNF data were collected retrospectively and had higher levels of missingness for select variables, which reduced analytic sample size and may have introduced bias. Fourth, the overall sample size constrained the number of predictors that could be included in multivariable modeling and limited statistical power to detect smaller effects. As such, findings should be interpreted with caution and validated in larger, multi-center samples. Finally, although we included established measures of stroke severity, comorbidity burden, and functional status, we did not capture all clinical or logistical factors that may influence discharge decisions. Variables such as medical complexity requiring specialized equipment (e.g., ventilatory support, dialysis, complex wound care), bed availability, or clinician assessment of medical stability were not available and may explain discharge decisions not fully captured by the variables available in this analysis.

## Conclusions

Among previously independent individuals with acute ischemic stroke and comparable measured clinical severity, discharge to IRF versus SNF was independently associated with non-clinical factors, particularly age and marital status, whereas functional status and comorbidity burden were not. These findings suggest that real-world post-acute rehabilitation placement may not consistently align with measurable indicators of rehabilitation need alone. More transparent and evidence-informed discharge frameworks may help better align rehabilitation intensity with recovery potential after stroke.

## Abbreviations

IRF: Inpatient Rehabilitation Facility
SNF: Skilled Nursing Facility
CCI: Charlson Comorbidity Index
LOS: Acute Care length of stay
AM-PAC: Activity Measure for Post-Acute Care
BM: Basic Mobility
DA: Daily Activity
AC: Applied Cognition
OR: Odds Ratio
CI: Confidence Intervals
PARC: Post-Acute Rehabilitation Care
NIHSS: National Institutes of Health Stroke Scale
PLOF: Prior Level of Function

## Data Availability

Available upon request

